# The Green Heart Project: Objectives, Design, and Methods

**DOI:** 10.1101/2023.12.05.23299461

**Authors:** Aruni Bhatnagar, Rachel Keith, Ray Yeager, Daniel Riggs, Clara Sears, Brent Bucknum, Ted Smith, Daniel Fleischer, Chris Chandler, Kandi L. Walker, Joy L. Hart, Sanjay Srivastava, Jay Turner, Shesh Rai

## Abstract

The Green Heart Project is a community-based trial to evaluate the effects of increasing greenery on urban environment and community health. The study was initiated in 2018 in a low-to-middle-income mixed-race residential area of nearly 28,000 residents in Louisville, KY. The 4 square mile area was surveyed for land use, population characteristics, and greenness, and assigned to 8 paired clusters of demographically- and environmentally matched “target” (T) and adjacent “control” (C), clusters. Ambient levels of ultrafine particles, ozone, oxides of nitrogen, and environmental noise were measured in each cluster. Individual-level data were acquired during in-person exams of 735 participants in Wave 1 (2018-2019) and 545 participants in Wave 2 (2021) to evaluate sociodemographic and psychosocial factors. Blood, urine, nail, and hair samples were collected to evaluate standard cardiovascular risk factors, inflammation, stress, and pollutant exposure. Cardiovascular function was assessed by measuring arterial stiffness and flow-mediated dilation. After completion of Wave 2, more than 8,000 mature, mostly evergreen, trees and shrubs were planted in the T clusters in 2022. Post planting environmental and individual-level data were collected during Wave 3 (2022) from 561 participants. We plan to continue following changes in area characteristics and participant health to evaluate the long-term impact of increasing urban greenery.

## INTRODUCTION

Globally, nearly 4.4 billion people live in urban areas, and this number is expected to increase to 6.6 billion by 2050.^4^ Although cities facilitate social interaction and provide economic opportunities, their high levels of pollution, noise, and crowding create adverse conditions that increase disease risk. Living in artificial urban environments also minimizes contact with nature, which seems conducive for psychological and physical health.^5^ Those who live in green areas report lower levels of stress^6–8^ and depression,^9–11^ whereas living in areas of low greenness is associated with increased risk of insulin resistance,^12^ diabetes,^13–15^ stroke,^16, 17^ as well as all cause, ^18^ ^19^ cancer,^20–22^ and cardiovascular mortality;^23, 24^ ^19^and decrease in life expectancy.^25^ Nevertheless, most of this evidence is observational and cross-sectional, which does not address causality or even the underlying mechanisms.

High levels of urban greenery could promote community health for many reasons.^24, 26–28^ Exposure to greenness could improve mental health and social cohesion. It could also promote physical activity and reduce exposure to noise, light, and air pollution. However, the extent to which these processes contribute to the health effects of greenery remains unclear. Moreover, because most epidemiological studies use satellite imagery (which only measures “greenness”),^29^ they provide little actionable data about the types, configurations, and locations of greenery that optimally benefit health. Therefore, we initiated the Green Heart Project (GHP) to directly examine changes in cardiovascular health and area characteristics in an urban neighborhood before and after a greening intervention in comparison with an adjacent area with no deliberate greening. We expect that interventional approach of the GHP would enable us to better address causal pathways linking greenness and health than the observational design of most studies to-date and provide key insights into mechanisms linking greenness and health.

A direct real-world, community-based evaluation of the link between greenness and cardiovascular health could inform the development of new approaches to prevent cardiovascular disease (CVD), which despite significant advances in treatment and management, remains the leading cause of death worldwide.^30^ Ominously, the rates of CVD continue to increase worldwide,^31^ in part due to increased urbanization as well as due to shifts in global temperature, increased pollution and extreme weather events due to climate change.^32^ Therefore, the results of GHP could help in assessing the efficacy of greenness as a unique, cost-effective public health strategy to decrease the global burden of CVD and to improve climate adaptation for people living in urban environments. Evaluation of the impact of urban greenness could also help in mitigating the effects of air pollution. Globally air pollution has been linked to 7 million premature deaths per year;^33^ 200,000 per year in the US alone.^34^ Because plants can remove pollutants,^35–48^ intensive greening could diminish air pollution particularly in areas of high roadway traffic. Increases in greenness could have other benefits as well, such as decreasing exposure to noise and light pollution, and buffering changes in ambient temperature. Thus, overall, the results of GHP could provide key insights into strategies for enhancing urban environments, improving public health, and mitigating the impact of climate change.

## MATERIAL AND METHODS

### Study Objectives

The objectives of GHP are to: 1) Determine the effect of a greening intervention on cardiovascular health of residents living in an urban neighborhood; 2) Quantify the impact of the greening intervention on neighborhood characteristics and air pollution and their contribution to cardiovascular health; 3) Assess the feasibility, sustainability, and suitability of the intervention; 4) Create a comprehensive database containing environmental, psychosocial, and clinical data to support long-term studies and data-sharing for additional ancillary studies; and 5) Develop an implementation framework to enhance health in urban neighborhoods by increasing area greenery.

### Overview of the Study Design

The GHP is a community-based quasi-experimental trial, with nonequivalent groups design,^49^ (National Clinical Trial number 03670524). The project tests the hypothesis that an increase in neighborhood greenery diminishes CVD risk by decreasing the levels of air pollution. To test this hypothesis, we collected baseline data on neighborhood characteristics and community health from 8 target (T) and 8 control (C) clusters of racially diverse, low-to-moderate income neighborhoods in Louisville, KY. We acquired aerial hyperspectral and Light Detection and Ranging (LiDAR) scans of the area and conducted stationary and mobile monitoring to develop high-resolution maps of ultrafine particles and select criteria air pollutants. We undertook extensive and frequent community engagement activities, and conducted in-person exams to acquire baseline physical and psychosocial data from >500 individuals from both C and T clusters. After acquiring baseline data, we planted >8000 mature trees and shrubs. Upon completion of planting, we evaluated changes in pollution and reacquired health data from in-person exams. To examine the effects of the greening intervention, we compare changes in CVD risk, psychosocial characteristics, and air pollution in C and T clusters. We will conduct follow up studies to determine how a progressive increase in the greenery will affect area characteristics and individual health in the neighborhood.

#### Selection and evaluation of residential clusters

To designate residential clusters for greening intervention, we evaluated existing tree cover, demographics, population density, and area traffic of all residential areas in Louisville, Kentucky, USA. We adopted a hierarchal approach for site selection to include areas with <30 % canopy, landcover suitable to implement an intervention to achieve substantially greater than 30% canopy and sufficient size and population density for enrollment of >700 study participants. Using this approach, we selected neighborhoods mostly within the 40214 zip code in south Louisville, although some areas extended slightly into other zip codes. The total area encompassed 4 sq. miles and has a population of 28,000 consisting of a little over 10,000 residential addresses. The median household income of the neighborhood is $32,600. The population is 54% White, 29% Black, 9% of 2 or more races, 8% of another race, and 11% Hispanic/Latino of any race; 32% of the population is 40-64 years old. To assess tree resources, we acquired aerial imagery of the area using a sub-meter multispectral and LiDAR sensors on a Cessna aircraft (Figure 1). In addition, certified arborists collected Global Positioning System (GPS) locations and the condition of 950 trees in the area. These ground truth points were used to train and validate LiDAR and hyperspectral data (Figure 1). For the study, the central zone of the area was divided into 8 clusters targeted for the greening intervention (Figure 2). To ensure geographic and demographic similarity, 8 clusters in the adjacent area, with greatest confluence of matching canopy cover, Normalized Difference Vegetation Index (NDVI), exposure to major roadways (I-264), total population, pollution exposure, household income, age, and ethnicity were designated as control (C).

**Figure. 1:**
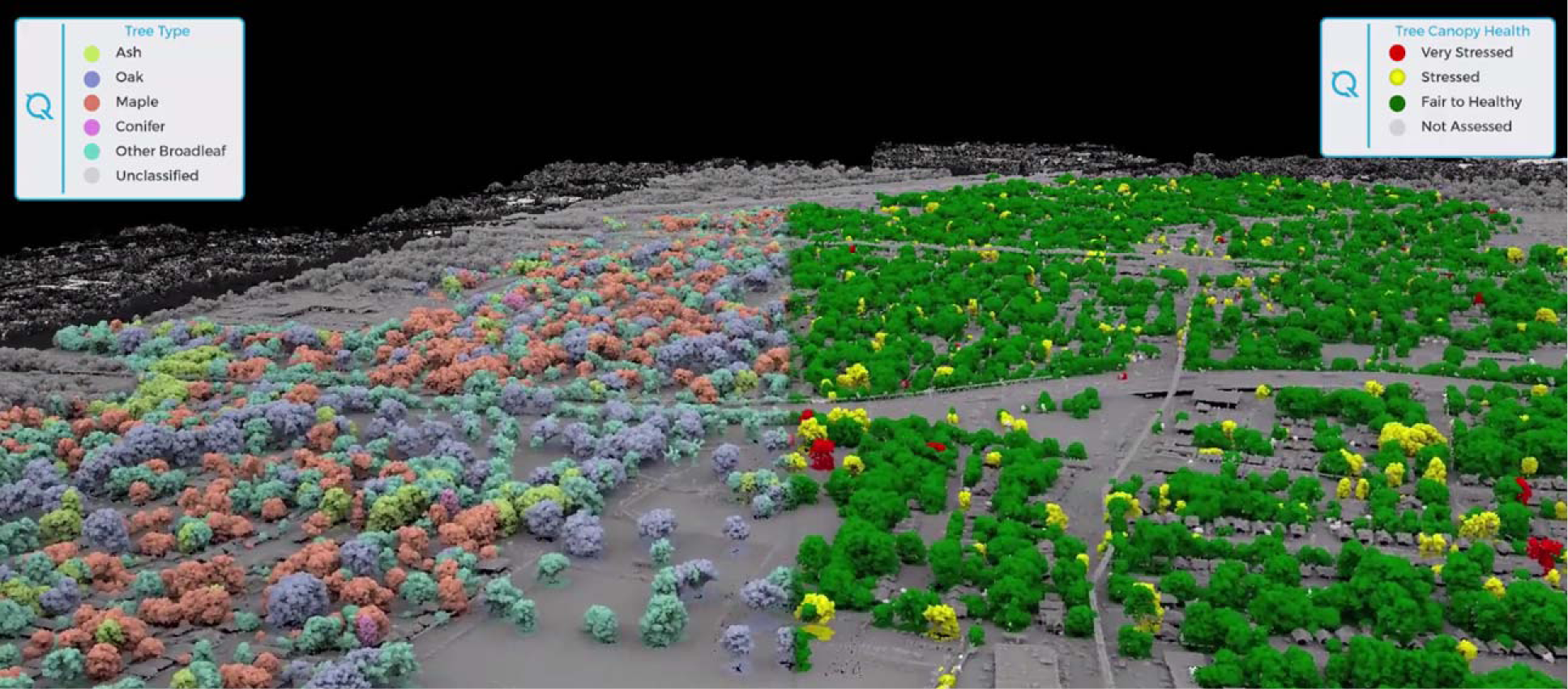
Hyperspectral scan of the Green Heart neighborhood

**Figure 2:**
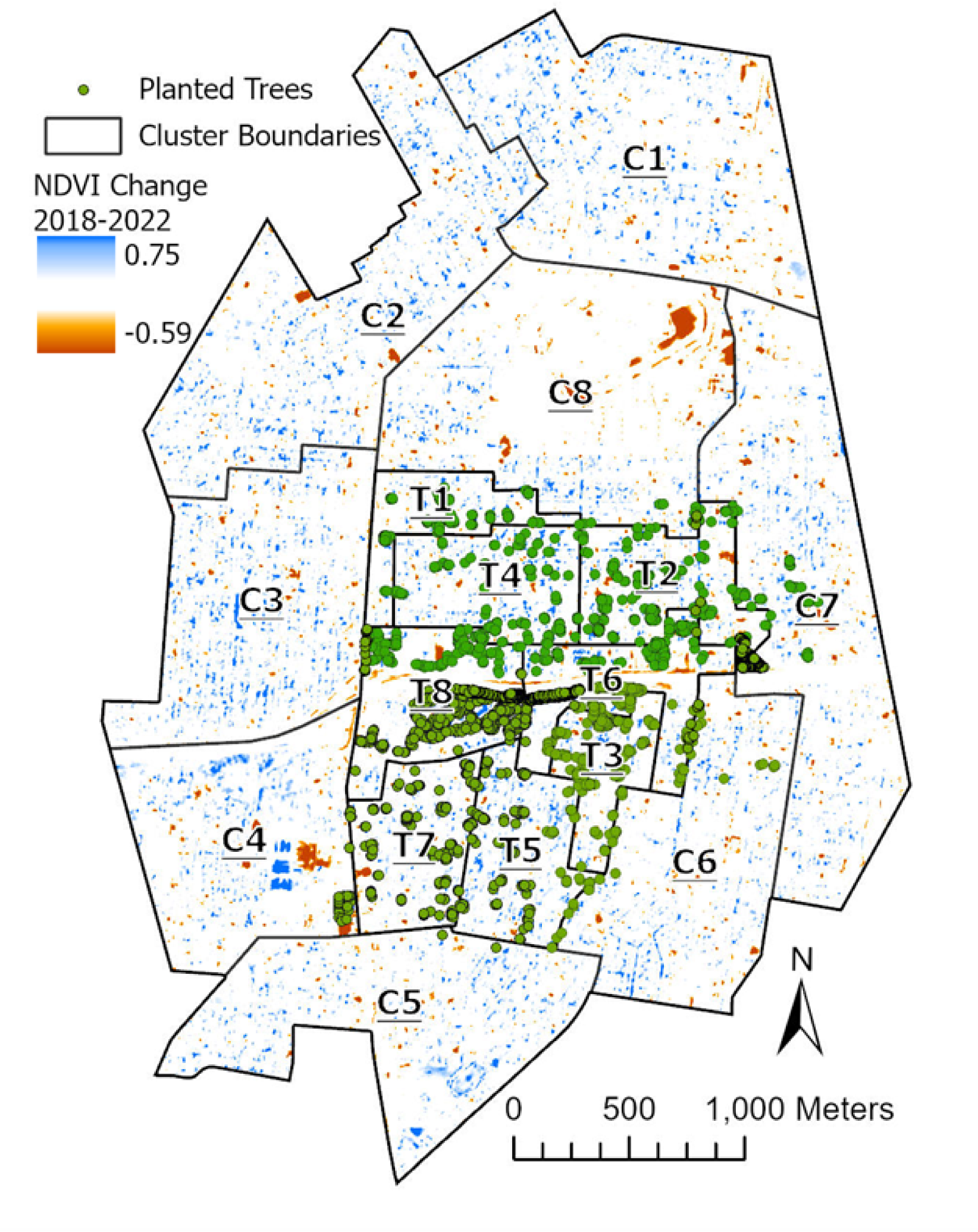
Target (T) and Control (C) neighborhood clusters within the study area. Areas in blue show an increase, whereas areas in red a decrease in NDVI. Planting areas are shown in green.

#### Community Engagement

Before finalizing the study area, we met with community leaders and residents to seek their input and participation. From conversations with community leaders and residents, we learned that there was interest in neighborhood improvements, connections between nature and health, and partnering with the university. Furthermore, in a recently completed neighborhood plan, preservation of mature trees and new plantings were identified as a neighborhood goal. We met with the Beechmont Neighborhood Association consisting of 10 board members and 5 community members, who were excited to participate in the project. We identified 50+ area businesses and organizations in the area and met representatives of each to seek their perception of the study. We engaged a local non-profit (Louisville Grows) to participate in our community engagement activities that included door-to-door canvasing. We participated and continue to participate in many community meetings and events each year as well as host opportunities for community member participation (e.g., health screenings, *Art and Literature Showcase*, *Health and Nature Photo Contests*, community presentations on environmental topics followed by question-and-answer sessions). Often, rather than holding separate GHP events, we support initiatives of community organizations, such as by volunteering and providing services. We have developed, and continue to develop, communication materials, including a website and regular updates, a quarterly newsletter, social media posts, infographics, and presentations for specific audiences, including schools and businesses. With this engagement and subsequent follow-up activities, we have developed an ongoing understanding of the community’s viewpoint and beliefs. Our conversations and relationships are ongoing, our communications are bi-directional, and learning occurs via multiple pathways. At the project’s inception, we formed a Community Advisory Board (CAB) populated by residents of the study area. The CAB meets quarterly and provides important insights and advice on GHP as well as community perspectives.

Community relationships are vital to the project. In a recent survey (Dec 22-Jan 23) of study participants and tree recipients (N=292), most respondents (89.7%) perceived GHP as a “great project.” Additionally, most (94.4%) believed that GHP trees would improve their health, and 92.8% indicated that GHP’s work enhances their community. Responses indicated increases in residents’ awareness of the effects of greenness and air quality on health (88.7% and 84.6%, respectively), the effects of the environment on heart disease (60.3%), the general positive aspects of spending time outdoors (68.5%), and the positive aspects of their community (60.3%). Moreover, most respondents (93.5%) perceived added trees as an improvement to the appearance of an area park. Notably, most (96.6%) hoped that GHP will continue working in their community.

#### Health Study Data Collection

Prior to the health study recruitment, we began to build our presence in the community and to foster awareness of the GHP. Curated recruitment materials were selected to develop cohesive branding and build awareness of the project. These materials were designed to specifically draw the attention of those interested in the environment, their neighborhood, and altruism. These cohesive and visually recognizable materials were used in a series of harmonized campaigns that included large community billboards and posters at bus stops, direct-to-resident mailers, in-person canvasing campaigns with door hangers, and digital imagery for social media placement. Each direct mail campaign for a wave of study visits consisted of a minimum of 3 touches for >10,000 residential addresses with a return of about 5-10% of those contacted completing study visits. Digital media campaigns garnered 50,000 impressions per week and click through rate of 0.16%. Our campaign also allowed us to engage in community events and meetings to attend, distribute GHP materials and to answer questions about the project. To supplement recruitment and community awareness, if they desired, enrolled participants were provided with approved recruitment materials to share with community members as well as yard signs to place at their residence with GHP branding and contact information. Retention efforts for returning participants included annual thank you events, birthday and holiday cards, community events, quarterly demographic updates as well as raffles for returning early in each wave. Returning participants received 3 touches by mail (either postcard or flyer) and 3 touches by email if they had a valid email address on file. Participants who did not respond to these methods of contact would be contacted by study staff via phone. We attempt a minimum of 4 unreturned phone calls prior to considering them a non-returner for that wave. Participants who reported moving out of the study area and still resided in Jefferson County, Kentucky, were allowed to continue with the study as an additional control population. All participants who never returned any method of contact for at least 2 waves or reported living outside of Jefferson County were not included in subsequent waves of visits.

#### Green Intervention

After the Wave 2 (2021) in-person exam, we planted trees and shrubs in T cluster neighborhoods (Figure 2). Because in comparison with deciduous trees, evergreen trees and shrubs are much more efficient at removing pollutants,^50–53^ and are green year-round, we planted mostly evergreens of 42 different species from 5 families (i.e., Pinaceae, Cupressaceae, Taxaceae, Aquifoliacae, and Magnoliaceae). We also planted some deciduous trees mainly Oak, Dogwood, and Serviceberry in residential areas and parks. To maximize biodiversity, we aimed to plant no more than 10% of the trees from the same species. Tree and shrubs were purchased from nurseries (in KY, OH, TN, IN, PA, and OR) and shipped to Louisville on trucks and temporarily stored in a staging area and spaded in at different sites. At baseline 26,256 trees existed in the study area, with 6,108 in T clusters and the rest in C clusters (Figure 2). At a cost of > 111,750-person hours, we planted 8,425 evergreen trees and shrubs and 630 deciduous trees in the T clusters, which more than doubled the number of trees in these areas. In addition, we treated 41 mature ash trees in the T clusters for infestation to prevent loss of tree canopy.

We sited 40% of planting in proximity to residences, while considering variables such as community and residential choices, soil characteristics, traffic density, codes, setbacks, signage visibility, and pedestrian safety. In addition, we conducted extensive plantings near roadways, mostly in the form of designed roadside barriers. We planted roadside trees in a multi-planting structure with both arbors and shrubs to enhance pollutant dispersal.^54^ Design parameters for roadside barriers were optimized to maximize pollution capture and dispersion with large trees in the back and smaller trees or shrubs in front. For each area, we targeted a wide distribution of species; considering sun exposure, soil type, disease, hydrology, community preference, aesthetics, and plant availability and suitability of the space for planting trees.

#### Measurements of Air pollution and Other Neighborhood Characteristics

We conducted and continue to conduct fixed site and mobile measurements of air pollution to estimate small-area variations. We use these data for LUR modeling to generate residence-level annual average ambient air pollutant concentrations. Passive samplers collect NO_x_, NO_2_, and O_3_ at 60 fixed sites using the location-allocation algorithm, with consistent temporal coverage (two weeks every two months) since 2018. Mobile platform measurements for ultrafine particle (UFP) number concentration, operationally defined as the concentration measured by a mixing condensation particle counter, are conducted by driving each public street and alley in the GHP study area multiple times over different times of day, days of week and season. Temperature monitors were added to the 60 passive sampler sites in November 2022. Noise was measured in collaboration with the Federal Aviation Administration (FAA) using high-quality Class 1 sound level meters. Four 2-to-3-week campaigns were conducted with 6-12 monitoring sites per campaign. To measure light pollution, we plan to collect data (5 years before and 5 years after planting) from the satellite-based Visible Infrared Imaging Radiometer Suite of instruments to measure the NDVI while accounting for non-vegetative influences on light from municipal GIS records.

#### In-person exams

To acquire health data, we invited residents to participate in the study at in-person exams conducted at study sites throughout the GH neighborhood from May to October. Participants were compensated $150 per visit. The study was approved by the University of Louisville IRB and all participants signed informed consent. We used 7 different sites that rotated and spanned the entire area to ease transportation and site-specific perception barriers. Study visits were scheduled over the course of all 7 days of the week with both daytime and evening hours to facilitate and maximize participation. These study visits were set up similar to a health fair allowing several participants per study visit that lasted several hours. There was a station set up for each measure which allowed participants to efficiently progress through the visit with less time and staff burden. During these in-person exams, participants were interviewed to obtain demographic data and to determine their medical history and lifestyle using validated questionnaires. Psychosocial factors were also assessed using validated questionnaires for stress,^55^ well-being,^56^ and social cohesion.^57^

We conducted the first wave of exams in 2018-2019 and then after a hiatus due to COVID-19 in 2020, we resumed study visits in 2021 (Figure 1). The format of the visits changed due to infection control protocols. Instead of rotating through the community, we stayed at an accessible location in the study area, and we invited participants to self-schedule appointments and complete surveys online prior to their in-person visit, reducing both staff and participant burden as well as contact time with individuals. If participants were not comfortable using our electronic system, we had dedicated study staff who answered phone calls and assisted participants in completing their surveys. Additionally, we began acquiring data on SARS-CoV-2 infection and vaccinations. In 2021, we recruited 545 participants, of which 416 were repeat visits, and we recruited 561 participants in 2022 of which 448 were repeat visits (Figure 3). Demographic characteristics of Wave 1 participants is shown in Table 1. We recruit individuals between 25-70 years of age, this will allow us to understand CV dysfunction due to pollutant exposure across the lifespan. ^58^

**Figure 3.**
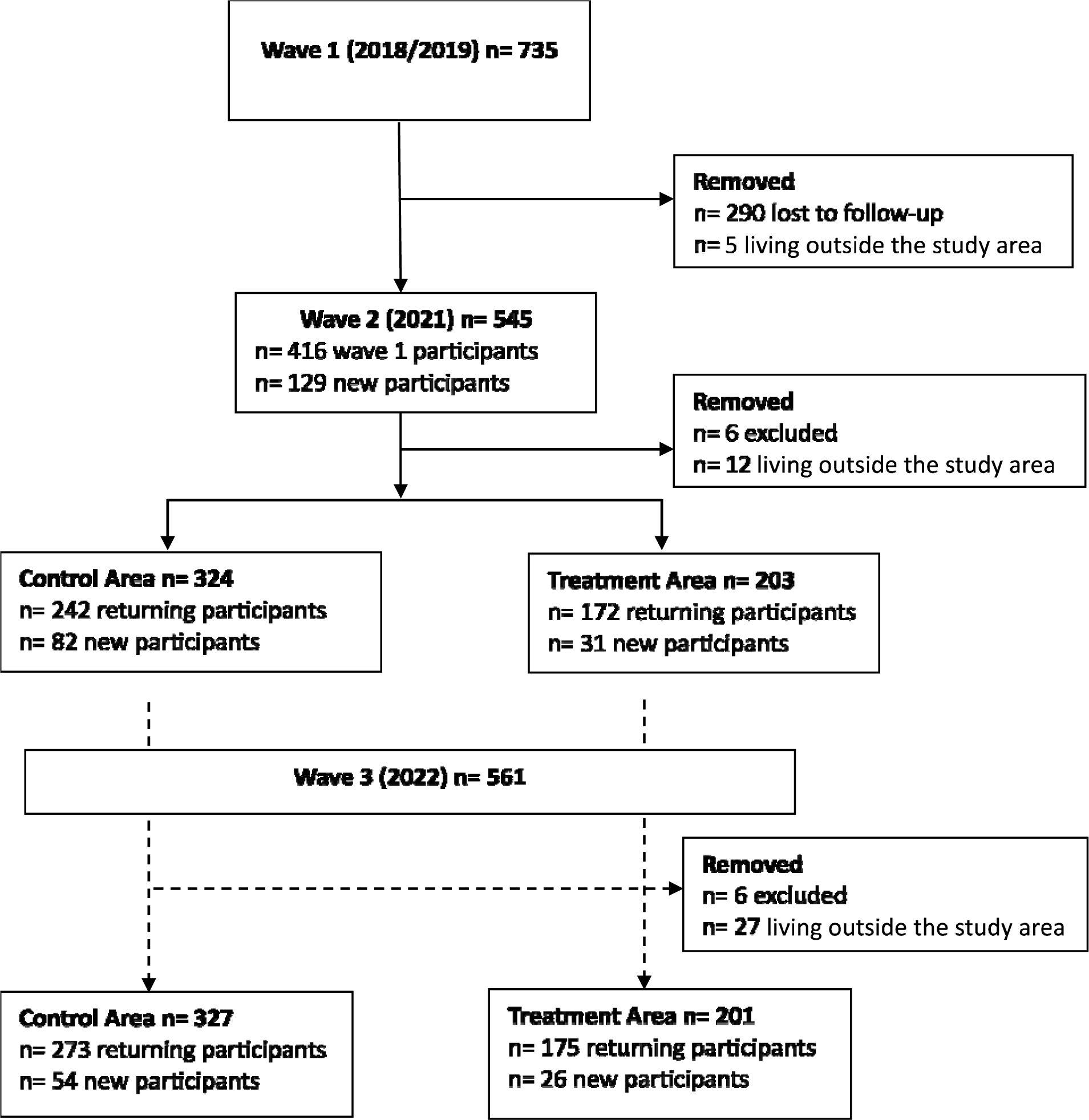
Flow Diagram of individuals who attended study visits and are eligible (Green Heart Study (2018-2022); Louisville, Kentucky). Solid and dashed lines indicate pre- and post-greening intervention, respectively. Eligible participants with study visits in all 3 Waves n=329 (Control Area n= 191; Treatment n=138).

**Table 1:**
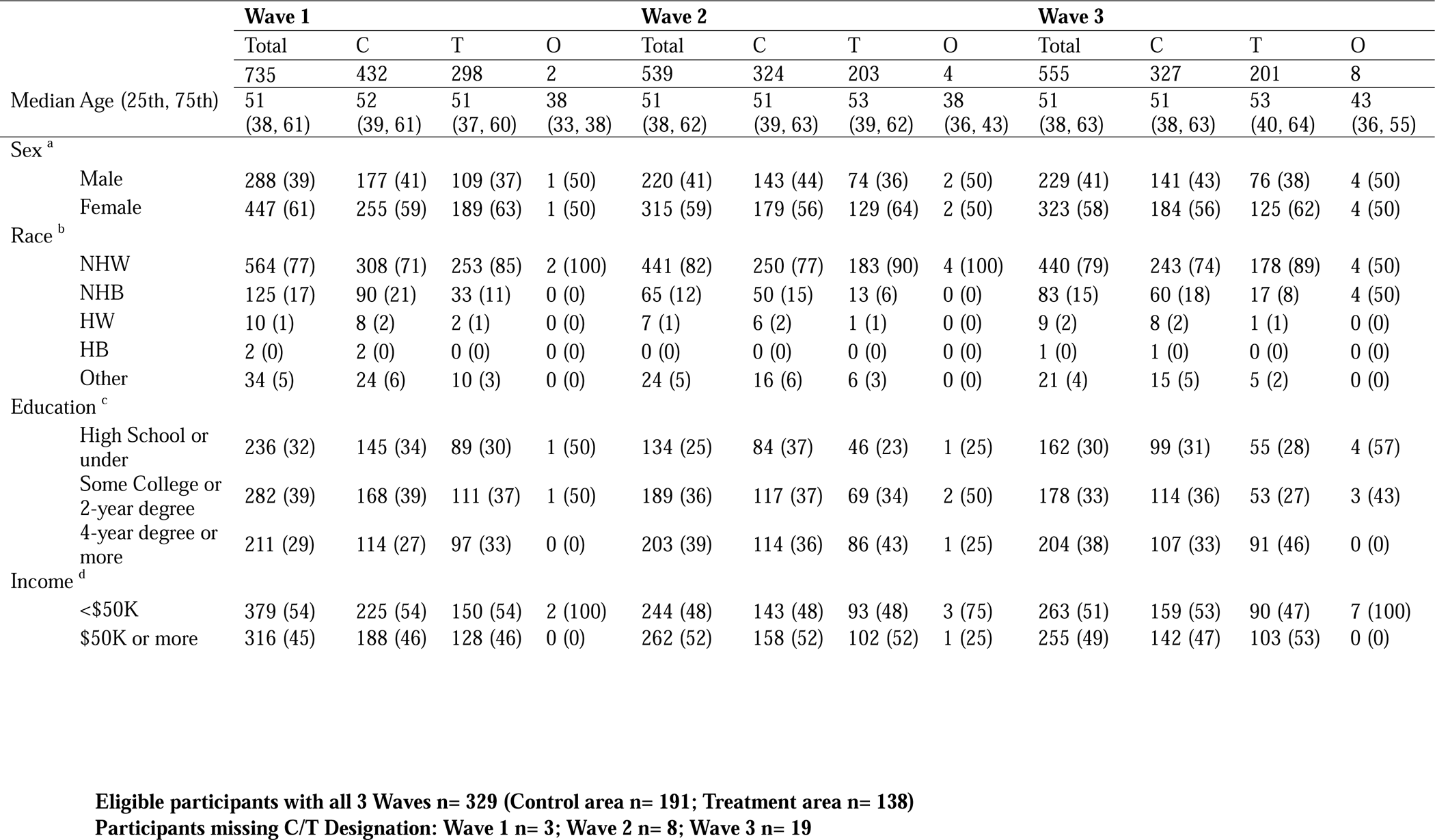
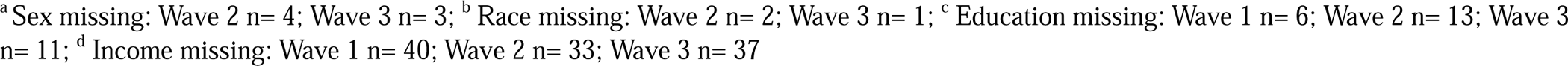
Demographic Characteristics of GHP participants.

During the in-person exam, we use standardized interviews to collect details about participants’ medical history, diet, exercise, and socioeconomic status using procedures similar to our previous Louisville Healthy Heart (LHHS) study (Table 2).^3, 59^ Psychosocial factors are assessed using validated questionnaires for stress,^60^, well-being,^61^ and social cohesion.^62^ Biological samples (blood, urine, nail, and hair) are obtained and stored. These are used for measuring biomarkers of exposure, stress^63, 64^ and CVD risk (Table 2). Trained research personnel use standardized procedure to measure height, weight, and waist and hip circumference (Table 3). To assess CVD risk, we acquire brachial blood pressure, arterial stiffness (augmentation index; AI, augmentation pressure, and pulse pressure), aortic systolic pressure and SEVR derived from pulse wave analysis using SphygmoCor, which provides highly reproducible values.^65–67^ Measures of arterial stiffness are positively and robustly associated with increased risk for CV events,^68^ and are associated with atherosclerosis, hypertension, stroke, Alzheimer-type dementia, and kidney disease. They provide novel and clinically-relevant information beyond that provided by standard risk factors; have been increasingly incorporated into longitudinal cohort studies.^68^ Measures of arterial stiffness complement blood pressure measurement but can also predict the onset of hypertension, cardiovascular mortality, incident cognitive decline and coronary artery disease.^68^ Previous work^69^ and our recent studies^70^ show that these measures are particularly sensitive to air pollution. Additionally, in a sub-set of participants, we measure flow mediated dilation as a measure of vascular health. Given the respiratory implications of air pollution we measure lung function by spirometry and oscillometeric methods. The in-person exam takes 45 min to complete, while surveys take between 45-90 min.

**Table 2:** Components of the In-Person Green Heart Examination.

|  | Description |
| --- | --- |
| **Information collected** |  |
| Personal Information | Name, date-of-birth, social security number, biologic sex (men or women) race (Black, White, other [American Indian/Alaskan Native, Asian, Hawaiian/Pacific Islander, other], age, education (high school or below, some college, college degree or higher) and annual household income (<\$20,000, \$20,000-\$44,999, \$45,000-\$64,999, ≥65,000) |
| Contact information | Residential address, list of household members |
| Informed consent | Obtain signed informed consent that complies with all required standards<br>Permission to contact for any future studies |
| Medical release | All study personnel to access participant's medical records |
| Medical history | General health status, cardiovascular disease history. Participants were asked to self-report their history of the following CVD conditions and events (coded as yes, no, do not know): stroke/transient ischemic attack, carotid endarterectomy or angiography, stenting/carotid angioplasty, heart attack/myocardial infarction, angina/stable coronary artery disease/atherosclerosis, heart valve disease, heart flutter/atrial fibrillation, percutaneous coronary intervention, and congestive heart failure. A composite variable, previous CVD, was also created, indexing whether or not participants had experienced any of the CVD events list above (coded as yes or no). For CVD risk participants reported on dyslipidemia/high cholesterol, diabetes and hypertension/high blood pressure (coded as yes, no, borderline, or do not know). |
| Medication use | Current use of medications for cardiovascular risk factor reduction and cardiovascular disease, analgesics, kidney or cancer drugs. |
| CVD risk | Framingham Risk Score was calculated using the following variables: sex (men or women), age (years), systolic blood pressure (millimeters of mercury), treatment for hypertension (yes or no), smoking (yes or no), diabetes (yes or no), high-density lipoprotein (HDL) (milligrams per deciliter), and total cholesterol (milligrams per deciliter). |
| BMI | Calculated from self-reported height and weight |
| Family history | Family's medical history of chronic disease |
| Occupational history | Specific occupation and aspects of occupation related to chronic disease and environmental exposures |
| Health care | Health insurance, use of health care facilities, barriers to health care and utilization access |
| Residential characteristics | Age of the house, house characteristics, attached car garage |
| Diet | Questions on dietary habits over the past 24 h plus a food propensity questionnaire |
| Smoking | Current smoker (yes or no), ever use of cigars and pipes, use of e-cigarettes and exposure to secondhand smoke.<br>Smoking status confirmed by urinary cotinine. |
| Drug use | Use of recreational drugs, including marijuana |
| Alcohol use | Alcohol use and duration |
| Exercise | Do you exercise regularly (>10 min each time)? Yes or no. |
| Perceived Stress | Global measurements of perceived stress.<sup>55</sup> |
| Well-being | Mental health was assessed using the Warwick Edinburgh Mental Well-Being Scale (WEMWBS) which is focused on positive aspects of mental health, including positive affect, fulfilling interpersonal relationships, and positive functioning.<sup>56</sup> |
| Depression | Depression status was evaluated based on responses to PHQ-9 items and not clinically diagnosed.<sup>75</sup> |
| Social cohesion | Social cohesion was assessed using a set of questions designed to capture individual residents' own level of integration in the neighborhood as well as the overall level of social cohesiveness they perceive to exist in the neighborhood. The questions also capture neighborhood physical and social disorder.<sup>57</sup> |
| Perception of Greenness | Individual perceptions of perceived greenness of neighborhood were measured using a scale comprising 15 perceived green items such as "lots of greenery around my local area, little," "no lawn nearby," or "walking or bicycle paths or trails nearby."<sup>76</sup> |
| Thought on nature and<br>Time spent in nature | View about nature, and daily time spent in nature |

**Table 3:**
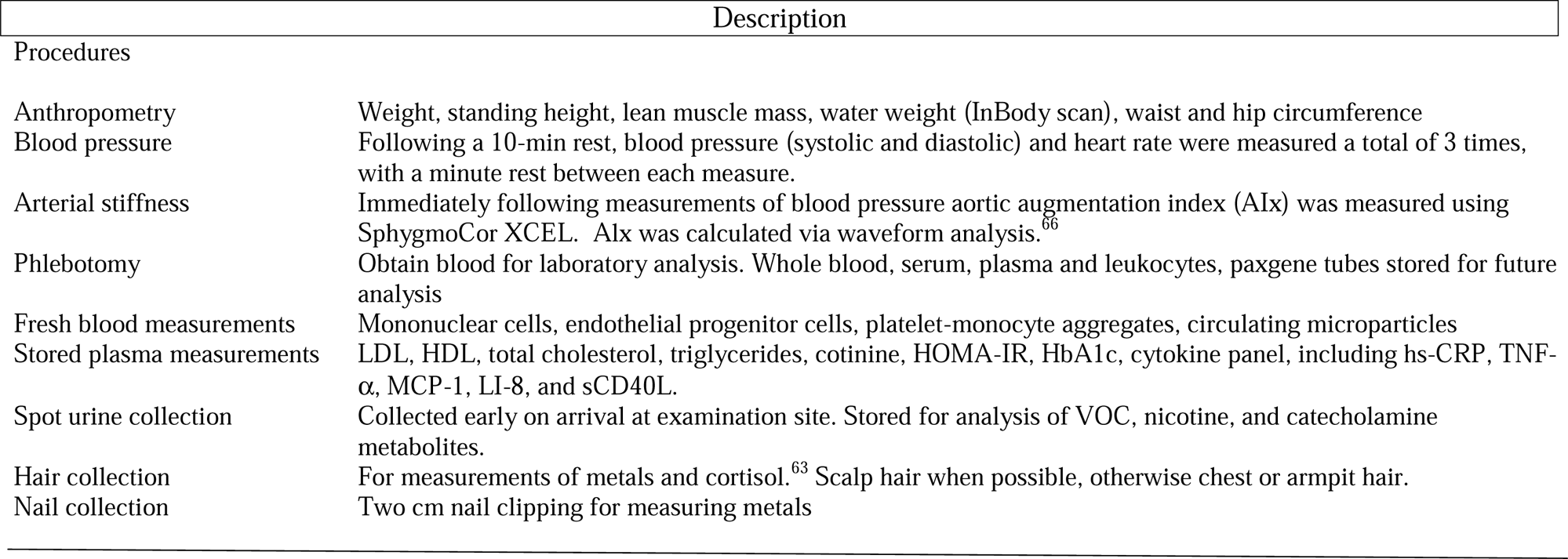
Exam Procedures.

#### Analysis Plan

To date, we have enrolled 964 unique participants who we will contact for post-intervention follow-up visits for Waves 4. We expect that there will be at least 200 (C) and 200 (T) participants after attrition. For longitudinal analysis, the sample size is justified in terms of our primary outcome – the augmentation index (AI). For sample size calculation, we used a two-sample one sided *t*-test for comparing the AI means in different groups. Due to one primary outcome variable, we set α=0.05 and with 90% power (β=0.20), we can detect an effect size of 0.24 SD (Table 4). To calculate power, we assumed a conservative positive correlation of 0.5 between two repeat observations. Furthermore, we can detect moderate to large effect sizes^71^ after adjusting for 3 covariates, which are moderately split (25-50%) in the population, using linear and logistic regression models.^72^ After pooling samples across multiple waves, we will have larger number of observations. Not only will we have at least 90% power to detect these differences in continuous outcomes and binary outcomes, we will also be able to adjust for multiple covariates and test for the contribution of demographic factors and mediators.

**Table 4:** Mean expected values and approximate effect sizes and power estimates for intra-individual changes in T clusters and population mean differences in C and T clusters. n_1_=250, n_2_=250.

| Outcomes | Value | Value | Effect size | Effect size | Intra-individual | Power | Power |
| --- | --- | --- | --- | --- | --- | --- | --- |
| Primary | Year 1 | Year 4 | Intra-individual | Population | Variation | Population | Intra-individual |
| AI | -24.3±8.6 | -28.9±10 | -25.8 | -30.9 | 5.2 | 90% | 95% |
| Secondary |  |  |  |  |  |  |  |
| Log PMA<sup>1</sup> | -2.22±0.6 | -2.22±0.6 | 7.7% | -9.24% | 0.04 | 90% | 95% |

We will use a pre-post quasi-experimental design (QED) with an equivalent control group to examine the effect of the greening. To assess balance, we will compare demographics, baseline health parameters, and neighborhood characteristics in C and T clusters. We will use linear mixed effects models (considering cluster as a random intercept effect) to examine the effect of the intervention on within-group and between-group differences in the primary (AI) and secondary individual health outcomes across all waves. Because we expect that the ‘dose’ of the greening intervention will increase over time, we will include an interaction between the in-person exam wave and cluster. These models will allow us to simultaneously account for the individual-level and cluster-level influences. To rule out historical bias and secular trends, we will compare T clusters with C clusters. The total effect of the intervention on individual health measures will be estimated at each wave as the difference in average change from baseline to wave, between C and T clusters. Models will be adjusted for demographic variables and other relevant confounders identified using a directed acyclic graph (DAG). The consistency of intervention effects on the primary outcomes will be explored in predefined subgroups that could influence an individual’s susceptibility to greening (i.e., time activity patterns), including sex, education, co-morbidities, and age categories. To strengthen internal validity, we will identify individuals with high and low exposures to investigate a “dose response” effect. As in our previous analyses of clinical trials, we will conduct case-controlled backward analysis dichotomizing participants that do or do not show an effect to identify potential non-responders and their characteristics.^73, 74^

#### Sustainability

To ensure sustainability, we will maintain the greening intervention. Newly planted trees, especially mature trees, require intensive care and watering for a minimum of the first two years. We will continue to water planted trees during summer (with more watering should there be <1 inch of rainfall over 10 days). Even with regular maintenance, we anticipate 8-11% mortality per year during the initial years, which we intend to monitor annually and remove and replace dead trees. To strengthen sustainability, we will continue close collaboration our CAB and area neighborhood associations and engage in regular community-wide presentations. We have contracted trained arborists to ensure that tree care needs are being met throughout the growing season. We have ongoing dialogue with the community to identify any issues that arise. For example, when the community raised concern regarding the cost of removing fallen or damaged trees, we set up a community fund to assist, even if these trees were not planted by us.

#### Data Dissemination and Data sharing

We will publish our findings in scientific peer-reviewed journals and present at scientific conferences. We will continue to engage with the lay press and share our experience with non-profit organizations such as The Nature Conservancy, urban planners, developers, and park services of different cities. We expect that greater visibility of the project will increase awareness of the value of greening and may promote implementation of similar interventions in cities worldwide. Moreover, several aspects of the project are ripe for collaboration in other areas than those currently included in GHP. These areas include studying the effects of greenness on asthma, sleep, respiratory disease, childhood development, birth outcomes, exercise, cancer, dementia, crime, economic status, storm water runoff, area biodiversity, heat island effect, energy consumption, and climate resilience. To enable studies in such diverse topics, we will deposit our data on the American Heart Association (AHA) Precision Medicine Platform. To gain access, investigators submit a brief manuscript proposal, which is reviewed by a manuscript committee (consisting of 6 investigators).

## DISCUSSION

The GHP will provide a rigorous and direct evaluation of the link between greenspaces and cardiovascular health. Findings of the project will help delineate the contribution of environmental, psychological, social, and physical factors to the beneficial effects of greenery and guide the development of healthy and sustainable urban neighborhoods. By evaluating the impact of greenness on pollution and temperature, the project could provide new insights into natural climate solutions. Lessons learned from implementation and sustainability, and the long-term cost-benefit analyses of the greening intervention could inform the development of large-scale environmental interventions to promote community health. Collectively, these results could inform new public health policies, better land use and development codes, tree ordinances, and facilitate evidence-based optimization of ongoing greening efforts worldwide. This information will be particularly critical as greening efforts worldwide are intensified to mitigate the adverse environmental effects of climate change.

Assessment of the longitudinal impact of the greening intervention will enable us to evaluate the long-term health effects of greening and how they may be related to changes in mental health and well-being and/or physical activity, standard cardiovascular risk factors, inflammation or stress. We also will be able to determine changes in physical activity and social cohesion and delineate the extent to which such behavioral changes contribute to the health effects of greenness. Our high-resolution mapping of area pollution will provide comprehensive information about how greenness affects pollutant levels and distribution, how specific patterns of planting alter pollution at specific sites, and how this distribution is related to health outcomes. To obtain a larger regional context, we will analyze our data within the context of regional data and the background levels of air pollution. Estimates of air pollution reduction vary significantly with different methodologies, assumptions about deposition rates and modeling approaches, and our analysis will provide comprehensive information about the extent of pollution reduction as a function of tree species, structure and placement. These results may provide new insights into the effects of urban vegetation on neighborhood temperature as well as noise and light pollution and will be directly relevant to the development of new public health policies and optimization of ongoing planting efforts in cities around the world to enhance public health and mitigate the effects of climate change. Collectively, the GHP will contribute to the formulation of an imitable paradigm, a model that could be readily replicated in other cities in both developed and developing countries – to reduce the levels of pollution, increase climate resilience and improve human health in urban environments.

## Data Availability

All non-protected data produced in the present study may be available upon reasonable request to the authors

## ACKNOWLEDGEMENTS

The Green Heart Project is supported by grants from the National Institutes of Environmental Health Sciences (ES 029846, ES030283) and The Nature Conservancy. We thank the community for their partnership and the Kentucky Transportation Cabinet and the Louisville Metro Government for providing access to public rights of way and staging spaces for the vegetation. We also acknowledge the in-kind support of the Federal Aviation Administration (FAA) and US DOT Volpe Transportation Research Center with equipment and personnel for noise monitoring. We also thank Dr. Jason Su, University of California at Berkeley for help in designing the passive sampling network, and Col. Blaine Hedges and his team at Government Solutions and Services for their help in planting trees.

